# Recovery and long-term health outcomes of SARS-CoV-2 infection in a prospective cohort in an urban setting, Kenya

**DOI:** 10.1101/2024.11.21.24316116

**Authors:** Isaac Kisiangani, Ângela Jornada Ben, Elke Wynberg, Welcome Wami, Samuel Iddi, Idah Kinya, Anna Vassall, Catherine Kyobutungi, Abdhalah Ziraba, John Njeru, Olive Mugenda, Marion Wangui Kiguoya-Njau, Mutambuki Kimondo, Geoffrey Githua, Menno D. de Jong, Shukri F. Mohamed, Gershim Asiki, Constance Schultsz

## Abstract

**Background:** Evidence on Long COVID remains limited in sub-Saharan countries. This study aimed to explore the occurrence of COVID-19-related symptoms and factors affecting recovery and Long COVID severity in Nairobi, Kenya.

**Methods:** A prospective cohort of individuals testing positive for SARS-CoV-2 between February 2022 and February 2023 were followed until June 2023. COVID-19-related symptoms were assessed every three months. Time to recovery was analyzed using survival analysis, while factors affecting recovery and Long COVID severity using Cox proportional hazard and Poisson regression, respectively.

**Results:** 42/291 (14%) participants had severe/critical SARS-CoV-2 infection, 59.1% were female and median age was 34. At 6 and 12 months post-positive PCR, 53.1% and 33.5% of participants had ≥1 COVID-19-related symptoms, respectively. Fatigue (40.2%), pain (36.8%), sore throat (36.8%), headaches (36.4%), and loss of strength (31.6%) were the most frequently reported COVID-19-related symptoms. Median time to recovery was longer in symptomatic participants with severe/critical SARS-CoV-2 infection than those with mild/moderate (234 vs 206 days respectively, p=0.016). Participants aged 40-64 years experienced slower recovery than those aged <40 years (aHR=0.635 [95%CI, 0.429;0.941]) and those with tertiary education recovered faster than those with primary education (aHR=1.869 [95%CI, 1.050;3.327]). Long COVID severity was associated with female sex (aIRR=1.413 [95%CI; 1.089;1.833]), tertiary education (aIRR, 0.525 [95%CI, 0.350;0.786]), and ≥1 comorbidity (aIRR=2.540 [95%CI, 1.377;4.687]).

**Conclusions:** Our findings suggest Long COVID presents a substantial, under-researched disease burden in Kenya. Risk factors for Long COVID are similar to those in high-income countries (HICs). Tailored prevention and support strategies for high-risk groups are needed.

## Introduction

The World Health Organization (WHO) estimates that 10 to 20% of individuals may experience persistent symptoms after a ‘Severe Acute Respiratory Syndrome Coronavirus-2’ (SARS-CoV-2) infection, also known as Post COVID-19 Condition or Long COVID [1]. Affected individuals include those who had initially mild symptoms or were asymptomatic at the time of initial SARS-CoV-2 infection [2,3]. Long COVID is defined by the WHO as the continuation or development of new symptoms three months after acute infection that last for at least two months and cannot be explained by an alternative cause [1,4]. Prevalence estimates of Long COVID widely vary due to differences in sample populations, study designs, and applied definitions of Long COVID [5]. Notably, data on Long COVID from low- and middle-income countries (LMICs), including sub-Saharan African countries, are scarce and few reported cohort studies had a short follow-up period [6]. A longitudinal study conducted in South Africa reported that 39% of participants had persistent COVID-19-related symptoms 6 months after infection, with fatigue being the most frequent symptom (32.1%) [7]. However, this study only followed patients for 6 months, hence the precise duration of post-COVID sequelae in these patients remained unclear. There is clearly an urgent unmet need for improved insights into the potential impact of Long COVID in sub-Saharan African countries.

In the current study, we aimed to investigate the occurrence of persistent COVID-19-related symptoms, potential factors influencing the time to recovery, and risk factors for Long COVID severity in a prospective cohort in the Nairobi Metropolitan region, the epicenter of the COVID-19 pandemic in Kenya. In addition, we specifically focused on factors associated with experiencing prolonged fatigue, representing the most commonly reported symptom of Long COVID. Finally, we assessed the quality of life of participants at 12-month of follow-up.

## Methods

This study is reported according to the Strengthening the Reporting of Observational Studies in Epidemiology (STROBE) Statement for cohort studies [8].

### Study design and enrolment

This is a prospective cohort study of individuals with a positive Polymerase Chain Reaction (PCR) result for SARS-CoV-2 at the Kenyatta University Teaching, Referral & Research Hospital (KUTRRH) and the Kenya Medical Research Institute (KEMRI). Eligible participants were enrolled between 8^th^ February 2022 and 21^st^ February 2023, and followed until 30^th^ June 2023. Individuals could join the study at any time after a positive PCR test (i.e. open enrolment). Inclusion criteria included age ≥18 years old, sufficient understanding of English or Kiswahili, and living in the Nairobi metropolitan area. Exclusion criteria were inability to comply with the study procedures as evaluated by the recruiting study team and living in long-term facilities.

The study team contacted individuals with a positive PCR test between December 2020 to July 2022 by telephone and distributed study flyers at the KUTRRH. Participants provided written informed consent at enrolment. This study was approved by the Kenya Medical Research Institute (KEMRI) Scientific and Ethics Review Unit (SERU – number 4306).

### Study procedures and data collection

Enrolment and follow-up took place at the KUTRRH. Participants were reimbursed for travel costs. Trained research assistants conducted study procedures and collected data via SurveyCTO tablets. Data collected at enrolment included age (years), sex, marital status, education level, occupation, socioeconomic status, self-reported health conditions (Asthma, Hypertension, Diabetes Mellitus, Dyslipidemia, Tuberculosis, HIV, Cancer), anthropometric measurements (height, weight), blood pressure, self-reported COVID-19 vaccination status, acute COVID-19 treatment (baricitinib, tocilizumab, remdesivir, dexamethasone), hospital admission, and oxygen therapy (Supplementary Text 1). The primary outcome was the presence of COVID-19-related symptoms measured using the identity section of the Illness Perception Questionnaire (IPQ-R) [9]. The IPQ-R identity section consists of 14 symptoms (any pain, sore throat, nausea, breathlessness, weight change, fatigue, stiff joints, sore eyes, wheeziness, headaches, upset stomach, sleep difficulties, dizziness, loss of strength). Participants were asked whether they experienced symptoms since their SARS-CoV-2 infection and whether they believed these symptoms were related to COVID-19 (Supplementary Methods 1). Responses were categorized as having no symptoms and having ≥1 COVID-19-related symptoms. Due to the absence of standardized criteria for assessing Long COVID severity, we assumed that a higher number of COVID-19-related symptoms corresponded to greater severity. The secondary outcomes were fatigue [10] and quality of life [11]. Fatigue was measured using the Fatigue Assessment Scale (FAS) and categorized as no fatigue (FAS score <22) and fatigue (FAS score ≥22) [10]. Quality of life was measured using the SF-36 comprising 36 questions that cover eight domains (i.e., general health, physical functioning, physical health, emotional health, social functioning, pain, energy/fatigue, and emotional well-being) [11]. The primary and secondary outcomes were measured at enrolment and every 3 months until 12 months, except for quality of life data collected at enrolment and 12 months only. A sample size of 168 symptomatic and 168 asymptomatic participants was estimated to detect a 20% difference in risk between mild/moderate and severe/critical SARS-CoV-2 infection severity (Supplementary Text 2).

### Statistical methods

Data were summarized as numbers and percentages for categorical variables, and as mean (SD) or median (IQR) for continuous variables, for the overall sample and by SARS-CoV-2 infection severity. Alluvial plots depicted the transitions between COVID-19-related symptoms over time and the frequency of individuals with new or relapsing symptoms. Incidence proportions of COVID-19-related symptoms since a positive PCR test were reported, representing the number of participants who ever reported symptoms among those still in follow-up at each time point since a positive PCR test.

Kaplan-Meier survival analysis estimated the proportion of participants experiencing ongoing COVID-19-related symptoms (i.e., not fully recovered), excluding asymptomatic participants at enrolment. Given participants were not enrolled in the study immediately after their positive PCR test, they became ‘at-risk’ upon enrolment (left-censored), with time measured in days since a positive PCR test. Thus, the at-risk period began on the positive PCR test day until recovery of COVID-19-related symptoms, loss-to-follow-up, death, or the last study visit, whichever came first. The analysis was stratified by SARS-CoV-2 infection severity. Potential factors influencing the time to recovery from COVID-19-related symptoms were explored using a univariable and multivariable Cox proportional hazard model. The choice of potential factors was based on the literature, including characteristics at enrolment (i.e., age, sex, marital status, education, employment status, occupation, socioeconomic status, comorbidities, and vaccination) [12]. Variables with a p-value <0.25 in the univariable analysis were included in the multivariable model to avoid overfitting and improve generalizability [13]. Unadjusted hazard ratio (HR) and adjusted hazard ratio (aHR) and their corresponding 95%CIs are presented. We assessed each HR using the proportional-hazards assumptions test based on Schoenfeld residuals and checked the validity of the proportional hazards assumption using stcoxkm [14] (Supplementary Methods 2).

Poisson generalized estimating equation (GEE) models were developed to assess risk factors associated with Long COVID severity [15]. GEE models included the total number of persistent COVID-19-related symptoms as an outcome and days since a positive PCR test, age, and sex as fixed covariates. Manual backward selection was used to retain variables in the model [13]. Variables included in the multivariable model were selected based on the p-value of <0.25 in the univariable analysis and clinical plausibility. Variables were included in the multivariable model in this order: sociodemographic and clinical characteristics; SARS-CoV-2 infection severity; and vaccination status. Adjusted incidence risk ratio (aIRR) and their 95%CIs are presented. We investigated interactions between BMI and other variables (i.e., comorbidity, age) as obesity clusters with other comorbidities [16].

A binomial GEE logistic model was conducted as a secondary analysis to assess the presence of fatigue over time using a similar development strategy as the Poisson GEE model. Results are presented as odds ratio (OR) and their 95%CIs. Fatigue was used as a proxy for Long COVID as fatigue is the most frequently reported persistent COVID-19-related symptom [17]. A linear regression was performed cross-sectionally to assess the association between Long COVID and quality of life at 12 months of follow-up, as suggested by existing literature [18]. All analyses were performed using Stata software (StataCorp, version 17.0).

## Results

### Study participants

A total of 2145 individuals were screened (Supplementary Figure S1), of whom 750 had a positive PCR test for SARS-CoV-2 infection. Of these, 459 individuals declined to participate (median age 43 (IQR=31-61), 54% female) and 291 participants were enrolled in the study (Supplementary Table 1a, Figure S1). The median age of participants was 34 (IQR=29-42) years and 172 (59.1%) were female (Table 1). The median time between a positive PCR test and enrolment into the study was 99 days (IQR= 71-169) with 113 of 291 (38.8%) participants enrolled less than 90 days after a positive PCR test. Five deaths occurred during follow-up (2 participants with mild/moderate and 3 participants with severe/critical SARS-CoV-2 infection) due to underlying comorbidity (HIV, diabetes, and hypertension complications).

**Table 1:** Characteristics of the participants of the Long COVID study by SARS-CoV-2 infection severity at diagnosis.

| **Characteristic** | **Overall**<br>**N=291** | **Mild/Moderate**<br>**N=249** | **Severe/Critical**<br>**N=42** |
| --- | --- | --- | --- |
| Age (median (IQR)) | 34 (29-42) | 32 (29-39) | 57 (40-63) |
| Age group (Years) |  |  |  |
| <40 | 198 (68.0) | 188 (75.5) | 10 (23.8) |
| 40-64 | 79 (27.1) | 57 (22.9) | 22 (52.4) |
| ≥65 | 14 (4.8) | 4 (1.6) | 10 (23.8) |
| Sex |  |  |  |
| Female | 172 (59.1) | 146 (58.6) | 26 (61.9) |
| Male | 119 (40.9) | 103 (41.4) | 16 (38.0) |
| Marital status |  |  |  |
| Married | 167 (57.4) | 140 (56.2) | 27 (64.3) |
| Never Married or Co-habited | 97 (33.3) | 92 (37.0) | 5 (11.9) |
| Divorced with a living partner | 18 (6.2) | 14 (5.6) | 4 (9.5) |
| Partner deceased | 9 (3.1) | 3 (1.2) | 6 (14.3) |
| Education level |  |  |  |
| None or Primary | 27 (9.3) | 18 (7.2) | 9 (21.4) |
| Secondary | 48 (16.5) | 36 (14.5) | 12 (28.6) |
| Tertiary | 216 (74.2) | 195 (78.3) | 21 (50.0) |
| Employed | 250 (85.9) | 225 (90.4) | 25 (59.5) |
| Self-reported socioeconomic status<sup>#</sup> |  |  |  |
| Low class | 32 (11.0) | 25 (10.0) | 7 (16.7) |
| Middle class | 158 (54.3) | 142 (58.0) | 16 (38.0) |
| Upper class | 68 (23.4) | 55 (22.1) | 13 (31.0) |
| Unknown | 33 (11.3) | 27 (10.9) | 6 (14.3) |
| Healthcare professionals | 112 (38.5) | 110 (44.2) | 2 (4.8) |
| BMI, Kg/M<sup>2</sup> |  |  |  |
| Normal weight (BMI <25 Kg/M<sup>2</sup>) | 126 (43.3) | 109 (43.8) | 17 (40.5) |
| Overweight (BMI 25-30 Kg/M<sup>2</sup>) | 95 (32.6) | 89 (35.7) | 6 (14.3) |
| Obesity (BMI >30 Kg/M<sup>2</sup>) | 70 (24.1) | 51 (20.5) | 19 (45.2) |
| Comorbidities |  |  |  |
| None | 219 (75.3) | 204 (81.9) | 15 (35.7) |
| One or more | 72 (24.7) | 45 (18.1) | 27 (64.3) |
| Asthma | 14 (4.8) | 11 (4.4) | 3 (7.1) |
| Hypertension | 37 (12.7) | 20 (8.0) | 17 (40.5) |
| Diabetes Mellitus | 17 (5.8) | 4 (1.6) | 13 (31.0) |
| Dyslipidaemia | 13 (4.5) | 8 (3.2) | 5 (11.9) |
| Tuberculosis | 9 (3.1) | 8 (3.2) | 1 (2.4) |
| HIV | 4 (1.4) | 3 (1.2) | 1 (2.4) |
| Cancer | 2 (0.7) | 1 (0.4) | 1 (2.4) |
| Symptom status at first study visit |  |  |  |
| Symptomatic | 187 (64.3) | 152 (61.0) | 35 (83.3) |
| Asymptomatic | 104 (35.7) | 97 (39.0) | 7 (16.7) |
| Fatigue at first study visit<sup>†*</sup> | 60 (20.6) | 43 (17.3) | 17 (40.5) |
| Good Quality of life (QoL) at first study visit<sup>\$</sup> | 246 (84.5) | 220 (88.3) | 26 (61.9) |
| COVID-19 vaccination prior to enrolment | 255 (87.6) | 226 (90.8) | 29 (69.1) |
| Covid-19 vaccine type |  |  |  |
| AstraZeneca | 197 (77.3) | 176 (77.8) | 21 (72.4) |
| Moderna | 28 (11.0) | 26 (11.5) | 2 (6.9) |
| Janssen J & J | 18 (7.0) | 13 (5.8) | 5 (17.2) |
| Pfizer | 12 (4.7) | 11 (4.9) | 1 (3.5) |
| Received medicine to treat COVID-19 | 33 (11.3) | 12 (4.8) | 21 (50.0) |
| Types of medicine to treat COVID-19 |  |  |  |
| Tocilizuma | 12 (4.1) | 1 (14.3) | 11 (52.4) |
| Remdesivir | 1 (0.3) | 0 (0.0) | 1 (2.4) |
| Dexamethasone | 15 (5.2) | 6 (2.4) | 9 (21.4) |
| Other long-term medication | 8 (2.7) | 6 (2.4) | 2 (4.8) |
| Admitted to the hospital and put on oxygen | 42 (14.4) | 0 (0.0) | 42 (100.0) |
| Home-based care and receiving oxygen | 6 (2.1) | 2 (0.8) | 4 (9.5) |
| Time from + PCR to first study visit (Median (IQR)) | 99 (71-169) | 99 (71-167) | 104 (75-171) |

IQR: Interquartile Range. Obesity: body mass index >30 Kg/M<sup>2</sup>. HIV: Human immunodeficiency virus. PCR: Polymerase chain reaction. QoL: Quality of life. SES: Socioeconomic status
Continuous variables are presented as median (IQR); categorical and binary variables are presented as N (%) and compared using the Pearson X<sup>2</sup> test.
Clinical severity groups are defined as follows: Mild/moderate COVID-19 infection was defined as participants reported not having had hospital admissions and oxygen therapy due to COVID-19 as well as home-based care and oxygen therapy and severe otherwise. Severe/critical COVID-19 infection was defined as participants reported having had hospital admission and/or needed oxygen therapy due to COVID
\*Fatigue: defined as no fatigue (FAS score <22) and Fatigue (FAS score score ≥ 22)
<sup>\$</sup>Quality of life (QoL): defined as poor QoL (overall score <50) and good QoL (overall score ≥50)
SES was measured using a ladder representing socioeconomic standing of people in the community. At the top were those who are best off, have most money, most education and most respected jobs. At the bottom are people who have least money and least jobs.
<sup>#</sup>The total did not up to 291 due to missing data.

### Incidence proportions of COVID-19-related symptoms

The most frequently reported COVID-19-related symptoms at enrolment were fatigue, pain, sore throat, headaches, and loss of strength at 40.2%, 36.8%, 36.8%, 36.4%, and 31.6%, respectively (Supplementary Table 1b; Supplementary Figure S2). At 3 months since a positive PCR test, fatigue (48.7%), pain (44.2%), headaches (44.2%), sore throat (40.7%), and loss of strength (40.7%) were still commonly reported symptoms. Fatigue, stiff joints, wheeziness, headaches, sleep difficulties, dizziness, and loss of strength were more frequently reported in the severe/critical infection group compared to the mild/moderate infection group (Table 2). The proportion of participants with at least one COVID-19-related symptom at 6, 12, 18, and 24 months since a positive PCR test was 53.1%, 33.5%, 39.7%, and 29.4%, respectively. Alluvial plots indicated that, while most of the participants transitioned to a lower number of symptoms over time, some developed new symptoms (Supplementary Figure S3).

**Table 2:** Incidence proportion of 14 COVID-19-related symptoms since a positive PCR by SARS-CoV-2 infection severity.

| Since positive PCR* | Month 3 |  |  |  | Month 6 |  |  |  | Month 9 |  |  |  | Month 12 |  |  |  |
| --- | --- | --- | --- | --- | --- | --- | --- | --- | --- | --- | --- | --- | --- | --- | --- | --- |
| Severity group | Overall | Mild/Mode rate | Severe/Critical | P-Value | Overall | Mild/Mode rate | Severe/Critical | P-Value | Overall | Mild/Moderate | Severe/Critical | P-Value | Overall | Mild/Mode rate | Severe/Critical | P-Value |
| N→<br>Symptom ↓ | N=113 | N=100 | N=13 |  | N=162 | N=137 | N=25 |  | N=197 | N=167 | N=30 |  | N=215 | N=186 | N=29 |  |
| Pain | 50 (44.2) | 46 (46.0) | 4 (30.8) | 0.298 | 51 (31.5) | 42 (30.7) | 9 (36.0) | 0.597 | 22 (11.2) | 15 (9.0) | 7 (23.3) | 0.022 | 28 (13.0) | 19 (10.2) | 9 (31.0) | 0.002 |
| Sore throat | 46 (40.7) | 42 (42.0) | 4 (30.8) | 0.438 | 52 (32.1) | 46 (33.6) | 6 (24.0) | 0.346 | 26 (13.2) | 21 (12.6) | 5 (16.7) | 0.542 | 29 (13.5) | 24 (12.9) | 5 (17.2) | 0.525 |
| Nausea | 23 (20.4) | 18 (18.0) | 5 (38.5) | 0.085 | 22 (13.6) | 18 (13.1) | 4 (16.0) | 0.701 | 15 (7.6) | 11 (6.6) | 4 (13.3) | 0.200 | 11 (5.1) | 8 (4.3) | 3 (10.3) | 0.169 |
| Breathlessness | 30 (26.5) | 23 (23.0) | 7 (53.9) | 0.018 | 36 (22.2) | 29 (21.2) | 7 (28.0) | 0.450 | 21 (10.7) | 14 (8.4) | 7 (23.3) | 0.015 | 17 (7.9) | 9 (4.8) | 8 (27.6) | <0.001 |
| Weight loss | 13 (11.5) | 9 (9.0) | 4 (30.8) | 0.021 | 21 (13.0) | 16 (11.7) | 5 (20.0) | 0.255 | 14 (7.1) | 9 (5.39) | 5 (16.7) | 0.027 | 6 (2.8) | 4 (2.2) | 2 (6.9) | 0.149 |
| Fatigue | 55 (48.7) | 47 (47.0) | 8 (61.5) | 0.324 | 51 (31.5) | 41 (29.9) | 10 (40.0) | 0.319 | 36 (18.3) | 25 (15.0) | 11 (36.7) | 0.005 | 27 (12.6) | 18 (9.7) | 9 (31.0) | 0.001 |
| Stiff joints | 28 (24.8) | 22 (22.0) | 6 (46.2) | 0.058 | 36 (22.2) | 28 (20.4) | 8 (32.0) | 0.201 | 23 (11.7) | 13 (7.8) | 10 (33.3) | <0.001 | 20 (9.3) | 14 (7.5) | 6 (20.7) | 0.023 |
| Sore eyes | 14 (12.4) | 13 (13.0) | 1 (7.7) | 0.585 | 21 (13.0) | 18 (13.1) | 3 (12.0) | 0.876 | 13 (6.6) | 12 (7.2) | 1 (3.3) | 0.434 | 12 (5.6) | 8 (4.3) | 4 (13.8) | 0.038 |
| Wheeziness | 18 (15.9) | 15 (15.0) | 3 (23.1) | 0.454 | 23 (14.2) | 18 (13.1) | 5 (20.0) | 0.366 | 17 (8.6) | 12 (7.2) | 5 (16.7) | 0.089 | 7 (3.3) | 3 (1.6) | 4 (13.8) | 0.001 |
| Headaches | 50 (44.2) | 42 (42.0) | 8 (61.5) | 0.182 | 52 (32.1) | 46 (33.6) | 6 (24.0) | 0.346 | 25 (12.7) | 20 (12.0) | 5 (16.7) | 0.477 | 28 (13.0) | 23 (12.4) | 5 (17.2) | 0.468 |
| Upset stomach | 16 (14.2) | 15 (15.0) | 1 (7.7) | 0.477 | 16 (9.9) | 11 (8.0) | 5 (20.0) | 0.065 | 6 (3.0) | 5 (3.0) | 1 (3.3) | 0.921 | 8 (3.7) | 5 (2.7) | 3 (10.3) | 0.043 |
| Sleep difficulties | 25 (22.1) | 22 (22.0) | 3 (23.1) | 0.930 | 33 (20.4) | 26 (19.0) | 7 (28.0) | 0.303 | 15 (7.6) | 8 (4.8) | 7 (23.3) | <0.001 | 15 (7.0) | 9 (4.8) | 6 (20.7) | 0.002 |
| Dizziness | 30 (26.5) | 24 (24.0) | 6 (46.2) | 0.089 | 26 (16.0) | 23 (16.8) | 3 (12.0) | 0.549 | 18 (9.1) | 11 (6.6) | 7 (23.3) | 0.003 | 15 (7.0) | 9 (4.8) | 6 (20.7) | 0.002 |
| Loss of strength | 46 (40.7) | 39 (39.0) | 7 (53.9) | 0.305 | 36 (22.2) | 27 (19.7) | 9 (36.0) | 0.072 | 25 (12.7) | 16 (9.6) | 9 (30.0) | 0.002 | 13 (6.0) | 9 (4.8) | 4 (13.8) | 0.060 |
| Overall symptoms | 82 (72.6) | 70 (70.0) | 12 (92.3) | 0.090 | 86 (53.1) | 69 (50.4) | 17 (68.0) | 0.104 | 70 (35.5) | 55 (32.9) | 15 (50.0) | 0.072 | 72 (33.5) | 55 (29.6) | 17 (58.6) | 0.002 |

\* The median time between a positive PCR and enrolment into the study was 99 days (IQR= 71-169) and 38.8% (n=113) out of 291 participants had less than 90 days between a positive PCR test and first visit.

**Table 2 continued**
| Since positive PCR | Month 15 |  |  |  | Month 18 |  |  |  | Month 21 |  |  |  | Month ≥24 |  |  |  |
| --- | --- | --- | --- | --- | --- | --- | --- | --- | --- | --- | --- | --- | --- | --- | --- | --- |
| Severity group | Overall | Mild/Moderate | Severe/Critical | P-Value | Overall | Mild/Moderate | Severe/Critical | P-Value | Overall | Mild/Moderate | Severe/Critical | P-Value | Overall | Mild/Moderate | Severe/Critical | P-Value |
| N→ | N=206 | N=179 | N=27 |  | N=194 | N=168 | N=26 |  | N=83 | N=73 | N=10 |  | N=34 | N=30 | N=4 |  |
| Symptom ↓ |  |  |  |  |  |  |  |  |  |  |  |  |  |  |  |  |
| Pain | 25 (12.1) | 19 (10.6) | 6 (22.2) | 0.085 | 35 (18.0) | 26 (15.5) | 9 (34.6) | 0.018 | 10 (12.0) | 9 (12.3) | 1 (10.0) | 0.832 | 5 (14.7) | 4 (13.3) | 1 (25.0) | 0.536 |
| Sore throat | 29 (14.1) | 23 (12.9) | 6 (22.2) | 0.192 | 36 (18.6) | 31 (18.5) | 5 (19.2) | 0.924 | 5 (6.0) | 4 (5.5) | 1 (10.0) | 0.573 | 3 (8.8) | 3 (10.0) | 0 (0.0) | 0.508 |
| Nausea | 6 (2.9) | 5 (2.8) | 1 (3.7) | 0.793 | 15 (7.7) | 11 (6.6) | 4 (15.4) | 0.116 | 3 (3.6) | 3 (4.1) | 0 (0.0) | 0.514 | 1 (2.9) | 1 (3.3) | 0 (0.0) | 0.711 |
| Breathlessness | 21 (10.2) | 14 (7.8) | 7 (25.9) | 0.004 | 27 (13.9) | 21 (12.5) | 6 (23.1) | 0.147 | 9 (10.8) | 8 (11.0) | 1 (10.0) | 0.927 | 1 (2.9) | 1 (3.3) | 0 (0.0) | 0.711 |
| Weight loss | 7 (3.4) | 6 (3.4) | 1 (3.7) | 0.925 | 16 (8.2) | 14 (8.3) | 2 (7.7) | 0.912 | 1 (1.2) | 1 (1.4) | 0 (0.0) | 0.710 | 0 (0.0) | 0 (0.0) | 0 (0.0) | - |
| Fatigue | 26 (12.6) | 21 (11.7) | 5 (18.5) | 0.322 | 41 (21.1) | 34 (20.2) | 7 (26.9) | 0.437 | 6 (7.2) | 6 (8.2) | 0 (0.0) | 0.347 | 5 (14.7) | 4 (13.3) | 1 (25.0) | 0.536 |
| Stiff joints | 11 (5.3) | 8 (4.5) | 3 (11.1) | 0.152 | 21 (10.8) | 17 (10.1) | 4 (15.4) | 0.421 | 3 (3.6) | 3 (4.1) | 0 (0.0) | 0.514 | 1 (2.9) | 0 (0.0) | 1 (25.0) | 0.005 |
| Sore eyes | 4 (1.9) | 2 (1.1) | 2 (7.4) | 0.027 | 14 (7.2) | 10 (6.0) | 4 (15.4) | 0.084 | 2 (2.4) | 2 (2.7) | 0 (0.0) | 0.596 | 1 (2.9) | 1 (3.3) | 0 (0.0) | 0.711 |
| Wheeziness | 6 (2.9) | 2 (1.1) | 4 (14.8) | <0.001 | 6 (3.1) | 5 (3.0) | 1 (3.9) | 0.812 | 2 (2.4) | 1 (1.4) | 1 (10.0) | 0.095 | 3 (8.8) | 1 (3.3) | 2 (50.0) | 0.002 |
| Headaches | 25 (12.1) | 22 (12.3) | 3 (11.1) | 0.861 | 35 (18.0) | 31 (18.5) | 4 (15.4) | 0.705 | 5 (6.0) | 5 (6.9) | 0 (0.0) | 0.393 | 3 (8.8) | 3 (10.0) | 0 (0.0) | 0.508 |
| Upset stomach | 6 (2.9) | 4 (2.2) | 2 (7.4) | 0.136 | 8 (4.1) | 6 (3.6) | 2 (7.7) | 0.325 | 3 (3.6) | 2 (2.7) | 1 (10.0) | 0.249 | 3 (8.8) | 3 (0.0) | 0 (0.0) | 0.508 |
| Sleep difficulties | 18 (8.7) | 12 (6.7) | 6 (22.2) | 0.008 | 18 (9.3) | 11 (6.6) | 7 (26.9) | 0.001 | 3 (3.6) | 3 (4.1) | 0 (0.0) | 0.514 | 2 (5.9) | 2 (6.7) | 0 (0.0) | 0.595 |
| Dizziness | 18 (8.7) | 12 (6.7) | 6 (22.2) | 0.008 | 18 (9.3) | 13 (7.7) | 5 (19.2) | 0.060 | 2 (2.4) | 2 (2.7) | 0 (0.0) | 0.596 | 3 (8.8) | 3 (10.0) | 0 (0.0) | 0.508 |
| Loss of strength | 18 (8.7) | 12 (6.7) | 6 (22.2) | 0.008 | 24 (12.4) | 18 (10.7) | 6 (23.1) | 0.075 | 3 (3.6) | 3 (4.1) | 0 (0.0) | 0.514 | 3 (8.8) | 2 (6.7) | 1 (25.0) | 0.225 |
| Overall symptoms | 65 (31.6) | 47 (26.3) | 18 (66.7) | <0.001 | 77 (39.7) | 61 (36.3) | 16 (61.5) | 0.014 | 18 (21.7) | 15 (20.6) | 3 (30.0) | 0.496 | 10 (29.4) | 8 (26.7) | 2 (50.0) | 0.336 |

### Time to recovery from COVID-19-related symptoms

A total of 171 participants were symptomatic at enrolment. Time to recovery was significantly longer in individuals with severe/critical acute COVID-19 than in those with mild/moderate COVID-19 (p=0.016, Log-rank test) (Figure 1). Among those with mild/moderate SARS-CoV-2 infection, the median time to complete recovery was 206 days (95%CI, 198; 239), while for those with severe/critical illness, the median time to recovery was 234 days (95%CI, 178; 487).

**Figure 1.**
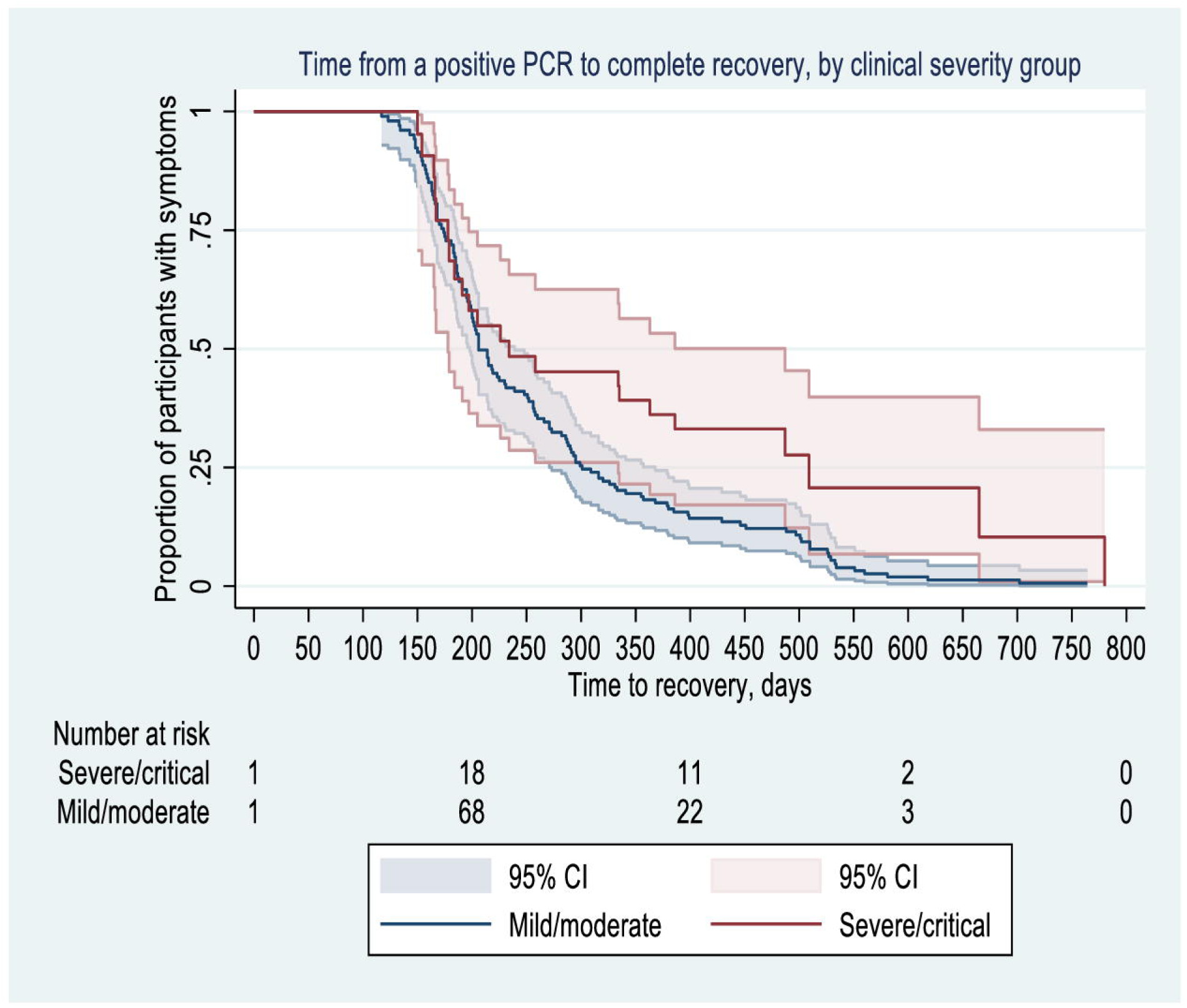
Kaplan Meier curve representing the proportion of participants with COVID-19-related symptoms (y-axis) in days between a positive PCR and complete recovery (x-axis), stratified by SARS­ CoV-2 infection (severe/critical *vs* mild/moderate). A total of 171 symptomatic participants were included in the survival analysis. As participants were not followed immediately after their positive PCR test, follow-up was left-censored, with participants becoming ‘at-risk’ when they were enrolled in the study. The curves represent the number of study participants recovering from COVID-19-related symptoms during the 2 years after a positive PCR. Shaded areas represent a 95% confidence interval (CI). The number of study participants at risk since positive PCR is below the graph. COVID-19: coronavirus disease 2019; SARS-CoV-2: Severe Acute Respiratory Syndrome Coronavirus 2.

### Factors influencing the time to recovery from COVID-19-related symptoms

There was no significant difference between sexes in time to recovery (aHR=0.787 [95%CI, 0.573; 1.082). Participants aged 40-64 years experienced 36.5% slower recovery than those aged <40 years (aHR=0.635 [95%CI, 0.429; 0.941]). Participants with tertiary education (aHR=1.869 [95%CI, 1.050; 3.327]) recovered almost twice as rapidly as those with primary education (Table 3).

**Table 3:** Cox proportional hazard model of factors influencing the time to recovery from COVID-19-related symptoms.

| Univariable Cox proportional hazard model |  |  | Multivariable Cox proportional hazard model |  |  |
| --- | --- | --- | --- | --- | --- |
| Factors | HR (95% CI) | p-value | Factors | aHR (95% CI) | p-value |
| **Sex** |  | 0.430 | **Sex** |  | 0.160 |
| Male | Ref |  | Male | Ref |  |
| Female | 0.888 (0.662 to 1.192) |  | Female | 0.787 (0.573 to 1.082) |  |
| **Age group (Years)** |  | 0.003 | **Age group (Years)** |  | 0.054 |
| <40 | Ref |  | <40 | Ref |  |
| 40-64 | 0.566 (0.385 to 0.832) |  | 40-64 | 0.635 (0.429 to 0.941) |  |
| ≥65 | 0.474 (0.185 to 1.213) |  | ≥65 | 0.687 (0.214 to 2.202) |  |
| **Education level** |  | 0.002 | **Education level** |  | 0.011 |
| Primary | Ref |  | Primary | Ref |  |
| Secondary | 1.244 (0.652 to 2.373) |  | Secondary | 1.258 (0.66 to 2.396) |  |
| Tertiary | 2.042 (1.16 to 3.593) |  | Tertiary | 1.869 (1.05 to 3.327) |  |
| **BMI, Kg/m2** |  | <0.001 | **BMI, Kg/m2** |  | 0.256 |
| Normal | Ref |  | Normal | Ref |  |
| Overweight | 1.252 (0.892 to 1.757) |  | Overweight | 1.097 (0.762 to 1.578) |  |
| Obese | 1.027 (0.697 to 1.512) |  | Obese | 1.211 (0.819 to 1.791) |  |
| **Comorbidity** |  | 0.057 | **Comorbidity** |  | 0.618 |
| None | Ref |  | None | Ref |  |
| One or more | 0.689 (0.469 to 1.012) |  | One or more | 0.877 (0.551 to 1.395) |  |
| **Vaccination status** |  | 0.046 | **Vaccination status** |  | 0.230 |
| Yes | Ref |  | Yes | Ref |  |
| No | 0.601 (0.365 to 0.99) |  | No | 0.734 (0.44 to 1.226) |  |

HR= Hazard ratio; aHR=Adjusted hazard ratio estimated using a multivariable Cox proportional hazard model including sex, age group, education, body mass index, comorbidities and vaccination status.

### Risk factors associated with Long COVID severity

Female participants were 1.4 times more likely than males to report a greater number of symptoms (aIRR=1.413 [95%CI, 1.089; 1.833]). Individuals with tertiary education were less likely to report a higher number of COVID-19-related symptoms compared to those with primary education (aIRR=0.525 [95%CI, 0.350; 0.786]). Having one or more comorbidities leads to a higher risk of reporting a greater number of COVID-19-related symptoms over time compared to having no comorbidity (aIRR=2.540 [95%CI, 1.377; 4.687]). On average, with each additional day since a positive PCR test, participants experienced a small but statistically significant reduction in the risk of reporting a greater number of COVID-19-related symptoms (aIRR=0.997 [95%CI, 0.996; 0.998]) (Table 4).

**Table 4:**
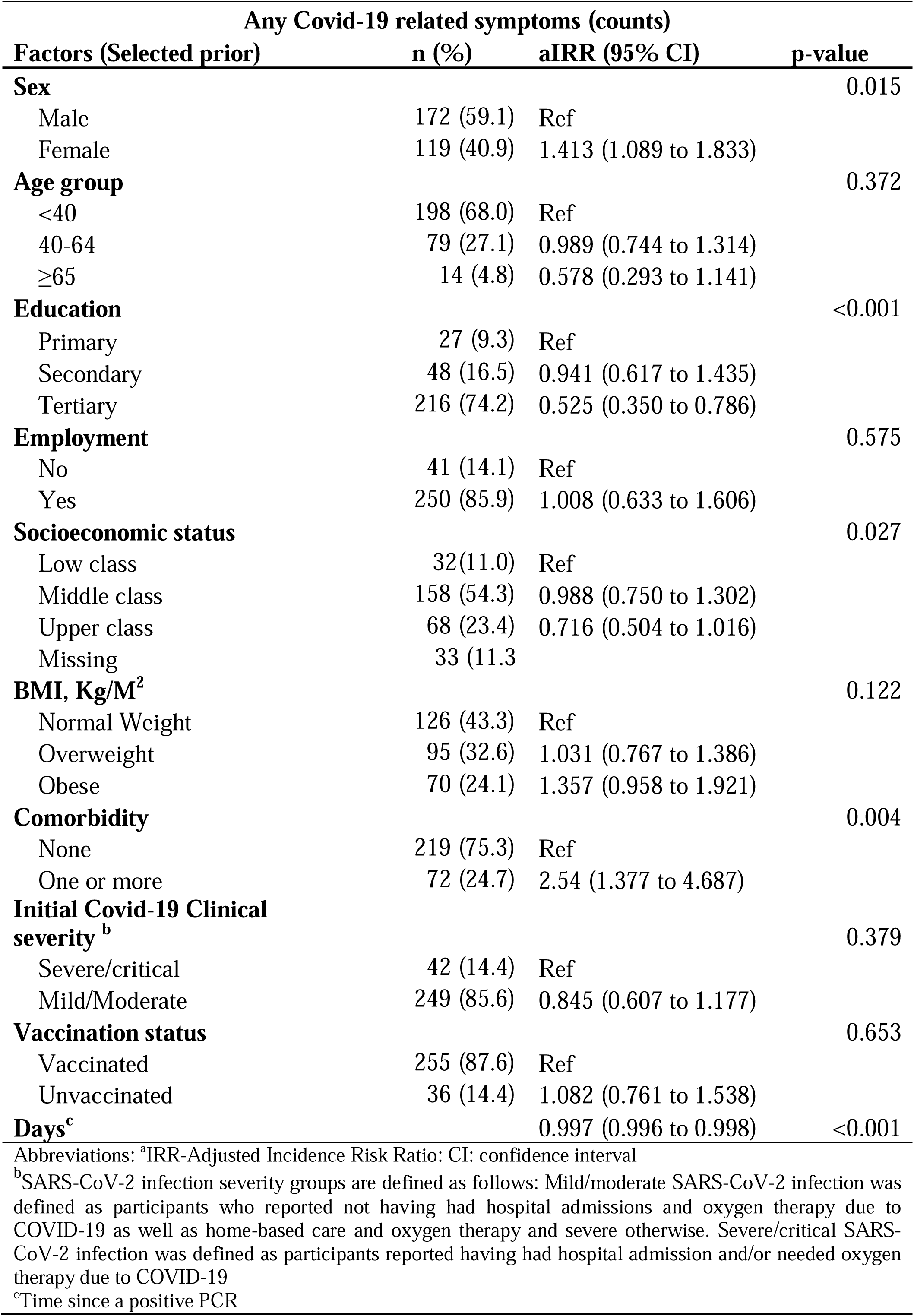
Poisson GEE multivariable model on risk factors associated with Long COVID severity over time.

### Fatigue and quality of life

Fatigue was 1.78 more likely to be reported by females than males (aOR=1.781 [95%CI, 1.128; 2.811]). The adjusted odds of reporting fatigue in employed individuals were about half (or 48% lower) than in unemployed individuals (aOR=0.518 [95%CI, 0.273; 0.981]). Individuals with high socioeconomic status were 50% less likely to have fatigue compared with those with low socioeconomic status (aOR=0.507 [95%CI, 0.316; 0.813]). Individuals with mild/moderate SARS-CoV-2 infection were 61% less likely to have fatigue compared to those with severe/critical (aOR=0.390 [95%CI, 0.199; 0.764]). On average, with an additional day since a positive PCR test, participants experienced a small but statistically significant decrease in risk for fatigue (aOR=0.999 [95%CI, 0.998; 1.000]) (Supplementary Table 2a).

Quality of life was significantly lower in individuals who experienced COVID-19-related symptoms at 12 months compared with those with no symptoms (aβ=-1.705 [95% CI, -2.595; -0.816]). Time since infection was not significantly associated with quality of life (p=0.326) when adjusting for other factors (Supplementary Table 2b).

## Discussion

At 6- and 12-months post-SARS-CoV-2-infection, half and one-third of participants, respectively, still reported COVID-19-related symptoms, with fatigue being the most commonly-reported complaint. These findings align with existing literature on Long COVID [19,20] . Older individuals (40-64 years) had slower recovery than those under 40, while participants with tertiary education recovered more quickly than those with primary education. Female participants, individuals with primary education, and those with one or more comorbidities were more likely to experience severe Long COVID.

Time to recovery from COVID-19-related symptoms was associated with older age, confirming other reports in the literature [21,22]. This may be due to age-related immune changes that increase susceptibility to infections and disease progression [23]. Due to the small number of participants aged 65 and older (n=14), we lacked the statistical power to determine if slower recovery applied to this age group. Highly educated participants were more likely to recover faster from COVID-19-related symptoms, aligning with a Ghanaian study that found individuals with tertiary education had better recovery rates than those with lower education [24].

When examining risk factors for Long COVID severity, we found that participants with tertiary education had a significantly lower risk of experiencing a higher number of symptoms compared with those with only primary education. This is consistent with findings from the literature as studies suggest that higher education predicts better health literacy, enabling easier access to healthcare [25,26]. High education often correlates with high socioeconomic status, enhancing the ability to take sick leave for recovery [27]. Indeed, in Kenya, higher education is typically associated with better socioeconomic status and comprehensive health insurance, easing medical costs, and improving access to healthcare [28,29]. This contrasts with Kenyan citizens who rely on the National Health Insurance Fund (NHIF) which, despite being the main health insurer, has limitations in coverage and quality of services [28,30]. Our findings highlight that higher education and socioeconomic status may directly and indirectly contribute to better outcomes such as recovery from COVID-19-related symptoms.

Consistent with reports from other settings, female sex was also significantly associated with Long COVID severity in our study [12]. Hypothesized mechanisms to explain the association between female sex and Long COVID include hormones perpetuating the hyperinflammatory status of the with the literature we found that having one or more comorbidities is associated with increased risk for Long COVID severity [7,33]. This could be due to immunosuppression caused by underlying health conditions, making them susceptible to severe infections and prolonged recovery periods [34]. In addition, many comorbidities are linked to chronic inflammation resulting in more severe inflammation [35]. However, we also recognize that participants may have reported symptoms related to pre-existing comorbidities during follow-up, and therefore we are uncertain to what extent the association between having comorbidities and severity of Long COVID is influenced by reporting bias.

In addition to exploring the duration and severity of Long COVID, we also specifically focused on fatigue, representing the most commonly reported Long COVID symptom. We found that post-COVID fatigue affects females more severely than males [36] and participants with severe SARS-CoV-2 infection reported more pronounced fatigue [37] than individuals with milder infections. Additionally, participants with higher socioeconomic status and those who are employed reported less fatigue compared to those from low socioeconomic backgrounds [37,38]. This is likely due to better access to healthcare, resources for recovery, and less exposure to stressors that can exacerbate fatigue, as noted previously. Finally, individuals with COVID-19-related symptoms were more likely to report poor QoL. This may be driven by the activity impairment, disability, and work-life disruption resulting from Long COVID, as previously implicated by other studies [39,40], and suggests a substantial psychosocial burden associated with Long COVID that may have profound socio-economic consequences if not addressed.

The comparison of results with other sub-Saharan countries was hampered due to limited research on Long COVID in African populations [6]. Interestingly, the overall risk factors identified in this study were similar to those found in high-income countries (HICs). Since the current study’s young, highly educated population (“healthy volunteer effect”) may indeed be comparable to populations studied in HICs. Further research is needed to understand Long COVID in more rural areas of Kenya and among lower socioeconomic groups with limited access to care.

Based on our knowledge, this study is the first in Kenya to report long-term health outcomes following a SARS-CoV-2 infection. However, the study had several limitations. The convenience sample introduced selection bias, hampering the generalization of results to the population with Long COVID in Kenya. SARS-CoV-2 PCR test costs were not covered by NHIF leading to a reduced sample size skewed towards highly-educated and middle-class individuals. Participants could join at any time since acute SARS-CoV-2 infection, with many enrolling ≥6 months after a positive PCR test, which introduces potential confounding. To address this, analyses were adjusted for time since the positive PCR test. The reliance on self-reported symptoms to define Long COVID may have also led to bias. The absence of a control group may limit the ability to account for possible confounders. Loss-to-follow-up affected 25% of the sample, despite mitigation strategies (i.e., phone reminders and transportation support).

## Conclusion

Six months after infection, half of the participants still had COVID-19-related symptoms, and a third continued to experience them after a year, with fatigue being the most common symptom. Faster recovery time from symptoms was associated with younger age and higher education level. Severe Long COVID was more likely in women and individuals with lower levels of education or pre-existing health conditions. Our findings underscore the potentially significant burden of Long COVID in Kenya, particularly in a more representative sample of the population, and the need to develop tailored support strategies for specific affected groups, particularly among vulnerable communities.

## Data Availability

All data produced in the present study are available upon reasonable request to the authors

## Acknowledgment

We are grateful to all of the study participants who took the time to contribute to our research as well as the field research team (Dorcas Keya, Catherine Kahare, Emmanuel Mwangi, Caliph Kirui, Ronald Makokite, Lilian Misati, Betty Chepkemoi and Benjamin Thoge) who helped collect data and Nelson Mbaya who was the data manager. We would also like to thank Dr. Antony Mulu of KUTRRH for his contributions to the project as consultant.

## Authors’ contributions

**IK:** Methodology, Validation, Data Curation, Formal analysis, Writing - Original Draft, Writing - Review & Editing; **ÂJB:** Methodology, Validation, Data Curation, Formal analysis, Writing - Original Draft, Writing - Review & Editing. **EW:** Data curation, Formal analysis, Writing-Review and Editing; **WW:** Conceptualization, Methodology, Validation, Data Curation, Formal analysis, Writing - Review & Editing. **SI:** Data curation, formal analysis, writing-review and editing; **IK:** Investigation, Review & Editing; **AV:** Conceptualization, Methodology, Resources, Funding acquisition, and Writing - Review & Editing; **CK:** Conceptualization, Writing - Review & Editing; **AZ:** Conceptualization, Writing - Review & Editing; **JN:** Conceptualization, Writing - Review & Editing; **OM:** Project administration; **MWK:** Conceptualization, Writing - Review & Editing; **MK**: Resources, project administration; **GG**: Resources, project administration; **MDJ:** Conceptualization, Methodology, Writing - Review & Editing; **GA:** Conceptualization, Methodology, Resources, Project administration, Funding acquisition, Writing - Review & Editing, Supervision. **CS:** Conceptualization, Methodology, Funding acquisition, Writing - Review & Editing.

