## Supplemental materials for "Recovery and long-term health outcomes of SARS-CoV-2 infection in a prospective cohort in an urban setting, Kenya"

**Supplementary files**

**Supplementary Text 1:** Definitions of study variables and outcomes

**Supplementary Text 2:** Sample size calculation

**Supplementary Table 1a: Age and sex characteristics of populations excluded and included from study**

**Supplementary Table 1b:** Incidence proportion of 14 COVID-19-related symptoms according to study follow-up periods by SARS-CoV-2 infection severity

**Supplementary Table 2a:** Binomial GEE multivariable model on factors associated with fatigue over time

**Supplementary Table 2b:** Linear regression of factors associated with Quality of life (QoL) at 12 months after the first visit

**Supplementary Figure S1:** Flow chart of participant enrolment into the cohort study.

**Supplementary Figure S2:** Prevalence of COVID-19-related symptoms at the first study visit (x-axis).

**Supplementary Figure S3:** Overview of data collection in the Long COVID-19 study, Nairobi Kenya

**Supplementary Figure S4:** Transition between levels of self-reported severity of Long COVID over time among prospectively included participants, clinical severity group

**Supplementary Methods 1:** Illness Perception Questionnaire (IPQ-R)

**Supplementary methods 2:** Test of the proportional-hazards assumptions after stcox using stcoxkm

**Supplementary Text 1:** Definitions of study variables and outcomes

COVID-19 illness onset was defined as the date of a positive PCR test obtained from the testing centres. Complete recovery was defined as the first moment upon which a participant reported no COVID-19-related symptoms in the past two weeks. Time to recovery was defined as the difference in days between the date of a positive PCR result (i.e., COVID-19 infection onset) and recovery from COVID-19-related symptoms. The severity of initial COVID-19 infection was categorized into mild/moderate and severe/critical, according to World Health Organization criteria [1]. Mild/moderate COVID-19 infection was defined as participants reported not having had hospital admissions and oxygen therapy due to COVID-19 as well as home-based care and oxygen therapy. Severe/critical COVID-19 infection was defined as participants reported having had hospital admission and/or needed oxygen therapy due to COVID-19. BMI was categorized as follows: underweight or normal weight (BMI <25 Kg/M^2^); overweight (BMI 25-30 Kg/M^2^), and obese (BMI >30 Kg/M^2^) (Tan, 2004, WHO 2000). A FAS score less than 22 indicates “normal” (i.e., healthy), between 22 and 34 indicates mild-moderate fatigue, and 35 or more indicates severe fatigue [2]. In this study, the FAS score was categorized as no fatigue (FAS score < 22) and fatigue (FAS score ≥ 22) as there was a low number of participants with severe fatigue (n=2). SF-36 scores range from 0 (worst) to 100 (best). Overall quality of life (QoL) scores were used to transform quality of life as poor QoL (overall QoL scores <50) and good QoL (overall QoL scores ≥50) [3]. Loss of follow-up was defined as withdrawal from the study or two consecutive unavailability despite 3 attempts to establish contact. Severity of Long COVID was defined as the total number of COVID-19-related symptoms; with a higher number of symptoms indicating more severe Long COVID.

**Supplementary Text 2:** Sample size calculation

Power calculations for various effect sizes were performed assuming that 50% of all participants diagnosed with COVID-19 would experience at least one symptom beyond three months from infection [4] and that one-third of these would be asymptomatic at diagnosis [5], a type 1 error of 𝛼=0.05, and a 95% confidence interval precision were used. A minimum sample of 129 symptomatic and 129 asymptomatic participants at diagnosis would be required to detect a difference of at least 20% (effect size, h=0.35) between the two groups (mild/moderate and severe/critical SARS-CoV-2 infection) with a power 80%, two-sided significance of 0.05 and using a Z- test. Assuming an estimated loss to follow-up of 30%, thus a total of 336 participants were to be recruited for follow-up.

**Supplementary Table 1a**: **Age and sex characteristics of populations excluded and included from study**

|  | Eligible | Refused to participate | Consented to participation | p-value |
| --- | --- | --- | --- | --- |
|  | N=750 | N =459**^†^** | N=291 |  |
| Sex (n, %) |  |  |  |  |
| Female | 420 (56.0) | 249 (54.2) | 172 (59.1) | 0.225 |
| Male | 330 (44.0) | 210 (45.8) | 119 (40.9) |  |
| Age, years (Median, IQR) | 38 (30-55) | 43 (31-61) | 34 (29-42) | 0.001 |
| **† Denotes the proportion of participants who refused to take part in the study after they were contacted and invited to take part in the study.** | | | | |

**Supplementary Table 1b: Incidence proportion of 14 COVID-19-related symptoms according to study follow-up periods by SARS-CoV-2 infection severity**

| **Follow-up period, months** | **first study visit*** | | | | | **3 months** | | | | | **6 months** | | | |
| --- | --- | --- | --- | --- | --- | --- | --- | --- | --- | --- | --- | --- | --- | --- |
| **Severity group** | **Overall** | **Mild/Moderate** | **Severe/ Critical** | **p-value** | **Overall** | | **Mild/Moderate** | **Severe/ Critical** | **p-value** | **Overall** | | **Mild/moderate** | **Severe/ Critical** | **p-value** |
| N→ | N=291 | N=249 | N=42 |  | N=251 | | N=218 | N=33 |  | N=229 | | N=198 | N=31 |  |
| Symptom ↓ |  |  |  |  |  |  |  |  |  |  |  |  |  |  |
| Pain | 107 (36.8) | 92 (37.0) | 15 (35.7) | 0.024 | 32 (12.7) | | 24 (11.0) | 8 (24.2) | 0.034 | 31 (13.5) | | 23 (11.6) | 8 (25.8) | 0.032 |
| Sore throat | 107 (36.8) | 93 (37.4) | 14 (33.3) | 0.620 | 40 (15.9) | | 36 (16.5) | 4 (12.1) | 0.52 | 26 (11.4) | | 21 (10.6) | 5 (16.1) | 0.367 |
| Nausea | 48 (16.5) | 35 (14.1) | 13 (31.0) | 0.006 | 21 (8.4) | | 19 (8.7) | 2 (6.1) | 0.608 | 5 (2.2) | | 4 (2.0) | 1 (3.2) | 0.669 |
| Breathlessness | 68 (23.4) | 51 (20.5) | 17 (40.5) | 0.005 | 28 (11.2) | | 23 (10.6) | 5 (15.2) | 0.434 | 17 (7.4) | | 9 (4.6) | 8 (25.8) | <0.001 |
| Weight loss | 41 (14.1) | 28 (11.2) | 13 (31.0) | 0.001 | 10 (4.0) | | 9 (4.1) | 1 (3.0) | 0.764 | 8 (3.5) | | 7 (3.5) | 1 (3.2) | 0.930 |
| Fatigue | 117 (40.2) | 93 (37.4) | 24 (57.1) | 0.016 | 43 (17.1) | | 33 (15.1) | 10 (20.3) | 0.031 | 26 (11.4) | | 20 (10.1) | 6 (19.4) | 0.131 |
| Stiff joints | 69 (23.7) | 49 (19.7) | 20 (47.6) | <0.001 | 22 (8.8) | | 17 (7.8) | 5 (15.2) | 0.164 | 19 (8.3) | | 13 (6.6) | 6 (19.4) | 0.016 |
| Sore eyes | 34 (11.7) | 29 (11.7) | 5 (11.9) | 0.962 | 21 (8.4) | | 18 (8.3) | 3 (9.1) | 0.872 | 10 (4.4) | | 6 (3.0) | 4 (12.9) | 0.012 |
| Wheeziness | 43 (14.8) | 32 (12.9) | 11 (26.2) | 0.024 | 18 (7.2) | | 15 (6.9) | 3 (9.1) | 0.647 | 8 (3.5) | | 4 (2.0) | 4 (12.9) | 0.002 |
| Headaches | 106 (36.4) | 89 (35.7) | 17 (40.5) | 0.555 | 37 (14.7) | | 30 (13.8) | 7 (21.2) | 0.261 | 26 (11.4) | | 24 (12.1) | 2 (6.5) | 0.355 |
| Upset Stomach | 36 (12.4) | 29 (11.7) | 7 (16.7) | 0.361 | 9 (3.6) | | 7 (3.2) | 2 (6.1) | 0.412 | 8 (3.5) | | 5 (2.5) | 3 (9.7) | 0.044 |
| Sleep difficulties | 62 (21.3) | 46 (18.5) | 16 (38.0) | 0.004 | 18 (7.2) | | 14 (6.4) | 4 (12.1) | 0.237 | 17 (7.4) | | 10 (5.1) | 7 (22.6) | 0.001 |
| Dizziness | 60 (20.6) | 48 (19.3) | 12 (28.6) | 0.168 | 19 (7.6) | | 15 (6.9) | 4 (12.1) | 0.289 | 17 (7.4) | | 11 (5.6) | 6 (19.4) | 0.006 |
| Loss of strength | 92 (31.6) | 71 (28.5) | 21 (50.0) | 0.006 | 27 (10.8) | | 21 (9.6) | 6 (18.2) | 0.14 | 11 (4.8) | | 8 (4.0) | 3 (9.7) | 0.172 |
| Overall symptoms | 187 (64.3) | 152 (61.0) | 35 (83.3) | 0.005 | 87 (34.7) | | 71 (32.6) | 16 (48.5) | 0.073 | 72 (31.7) | | 53 (26.8) | 19 (61.3) | 0.000 |
| ***** The median time between a positive PCR test and enrolment into the study was 99 days (IQR= 71-169) and 38.8% (n=113) out of 291 participants had less than 90 days between a positive PCR test and first visit. | | | | | | | | | | | | | | |

**Supplementary Table 1b continued**

| **Follow-up period, months** | **9 months** | | | | **12 months** | | | |
| --- | --- | --- | --- | --- | --- | --- | --- | --- |
| **Severity group** | **Overall** | **Mild/Moderate** | **Severe/ Critical** | **p-** | **Overall** | **Mild/Moderate** | **Severe/ Critical** | **p-value** |
|  |  |  |  | **value** |  |  |  |  |
| N→ | N=216 | N=188 | N=28 |  | N=219 | N=189 | N=30 |  |
| Symptom ↓ |  |  |  |  |  |  |  |  |
| Pain | 22 (10.2) | 17 (9.0) | 5 (17.9) | 0.15 | 34 (15.5) | 25 (13.2) | 10 (33.3) | 0.005 |
| Sore throat | 23 (10.6) | 17 (9.0) | 6 (21.4) | 0.035 | 30 (13.7) | 27 (14.3) | 3 (10.0) | 0.526 |
| Nausea | 6 (2.8) | 5 (2.7) | 1 (3.6) | 0.784 | 16 (7.3) | 12 (6.4) | 4 (13.3) | 0.172 |
| Breathlessness | 23 (10.6) | 15 (8.0) | 8 (28.6) | 0.001 | 26 (11.9) | 21 (11.1) | 5 (16.7) | 0.382 |
| Weight loss | 10 (4.6) | 8 (4.3) | 2 (7.1) | 0.498 | 9 (4.1) | 7 (3.7) | 2 (6.7) | 0.448 |
| Fatigue | 24 (11.1) | 21 (11.2) | 3 (10.7) | 0.943 | 37 (16.9) | 29 (15.3) | 8 (26.7) | 0.124 |
| Stiff joints | 13 (6.0) | 9 (4.8) | 4 (14.3) | 0.049 | 20 (9.1) | 17 (9.0) | 3 (10.0) | 0.859 |
| Sore eyes | 9 (4.2) | 8 (4.3) | 1 (3.6) | 0.866 | 7 (3.2) | 5 (2.7) | 2 (6.7) | 0.245 |
| Wheeziness | 8 (3.7) | 3 (1.6) | 5 (17.9) | <0.001 | 5 (2.3) | 3 (1.6) | 2 (6.7) | 0.084 |
| Headaches | 25 (11.6) | 23 (12.2) | 2 (7.1) | 0.432 | 29 (13.2) | 26 (13.8) | 3 (10.0) | 0.573 |
| Upset Stomach | 2 (0.9) | 2 (1.1) | 0 (0.0) | 0.583 | 11 (5.0) | 8 (4.2) | 3 (10.0) | 0.176 |
| Sleep difficulties | 14 (6.5) | 11 (5.9) | 3 (10.7) | 0.329 | 18 (8.2) | 12 (6.3) | 6 (20.0) | 0.011 |
| Dizziness | 14 (6.5) | 9 (4.8) | 5 (17.9) | 0.009 | 20 (9.1) | 14 (7.4) | 6 (20.0) | 0.026 |
| Loss of strength | 16 (7.4) | 10 (5.3) | 6 (21.4) | 0.002 | 22 (10.0) | 16 (8.5) | 6 (20.0) | 0.051 |
| Overall symptoms | 66 (30.6) | 51 (27.1) | 15 (53.6) | 0.005 | 68 (31.1) | 53 (28.0) | 15 (50.0) | 0.016 |

**Supplementary Table 2a: Binomial GEE multivariable model on factors associated with fatigue over time**

| **Fatigue** | | | |
| --- | --- | --- | --- |
| **Factors (Selected prior)** | **n (%)** | **aOR (95% CI)** | **p-value** |
| **Sex** |  |  | 0.006 |
| Male | 172 (59.1) | Ref |  |
| Female | 119 (40.9) | 1.781 (1.128 to 2.811) |  |
| **Age group** |  |  | 0.735 |
| <40 | 198 (68.0) | Ref |  |
| 40-64 | 79 (27.1) | 0.999 (0.596 to 1.674) |  |
| ≥65 | 14 (4.8) | 0.843 (0.266 to 2.672) |  |
| **Education level** |  |  | 0.515 |
| Primary | 27 (9.3) | Ref |  |
| Secondary | 48 (16.5) | 0.677 (0.313 to 1.464) |  |
| Tertiary | 216 (74.2) | 0.699 (0.354 to 1.381) |  |
| **Socioeconomic status** |  |  | 0.001 |
| Low class | 32(11.0) | Ref |  |
| Middle class | 158 (54.3) | 0.977 (0.678 to 1.408) |  |
| Upper class | 68 (23.4) | 0.507 (0.316 to 0.813) |  |
| Missing | 33 (11.3 |  |  |
| **Employment** |  |  | 0.047 |
| No | 41 (14.1) | Ref |  |
| Yes | 250 (85.9) | 0.518 (0.273 to 0.981) |  |
| **BMI, Kg/M^2^** |  |  | 0.767 |
| Normal Weight | 126 (43.3) | Ref |  |
| Overweight | 95 (32.6) | 1.159 (0.686 to 1.958) |  |
| Obese | 70 (24.1) | 0.9 (0.510 to 1.586) |  |
| **Comorbidity** |  |  | 0.743 |
| None | 219 (75.3) | Ref |  |
| One or more | 72 (24.7) | 1.1 (0.623 to 1.94) |  |
| **Covid-19 Clinical severity ^b^** |  |  | 0.005 |
| Severe/critical | 42 (14.4) | Ref |  |
| Mild/Moderate | 249 (85.6) | 0.39 (0.199 to 0.764) |  |
| **Vaccination status** |  |  | 0.566 |
| Vaccinated | 255 (87.6) | Ref |  |
| Unvaccinated | 36 (14.4) | 1.225 (0.651 to 2.302) |  |
| **Days^c^** |  | 0.999 (0.998 to 1) | 0.010 |
| aOR: Adjusted Odds ratio, CI: Confidence Interval. Fatigue was measured using fatigue assessment scale (FAS) and Fatigue: defined as no fatigue (FAS score <22) and Fatigue (FAS score score ≥ 22). Clinical severity groups are defined as follows: Mild/moderate SARS-CoV-2 infection was defined as participants who reported not having had hospital admissions and oxygen therapy due to COVID-19 as well as home-based care and oxygen therapy and severe otherwise. Severe/critical SARS-CoV-2 infection was defined as participants reported having had hospital admission and/or needed oxygen therapy due to COVID-19.  ^c^Time since positive PCR | | | |

**Supplementary Table 2b: Linear regression of factors associated with Quality of life (QoL) at 12 months after the first visit**

| **Quality of life at 12 months after first visit** | | |
| --- | --- | --- |
| **Factors** | **β coef. (95% CI)** | **p-value** |
| Sex |  | 0.051 |
| Male | Ref |  |
| Female | -2.166 (-5.112 to 0.779) |  |
| Age group (years) |  | 0.029 |
| <40 | Ref |  |
| 40-65 | -2.116 (-5.945 to 1.712) |  |
| >65 | -11.622 (-22.451 to -0.793) |  |
| Education |  |  |
| Primary | Ref | 0.650 |
| Secondary | 6.984 (-1.509 to 15.488) |  |
| Tertiary | 4.005 (-3.682 to 11.691) |  |
| **Socioeconomic status** |  | <0.001 |
| Low class | Ref |  |
| Middle class | 5.286 (-0.822 to 11.394) |  |
| Upper class | 11.165 (4.764 to 17.567) |  |
| Missing |  |  |
| BMI (KG/M2) |  | 0.732 |
| Normal | Ref |  |
| Overweight | 0.417 (-2.899 to 3.733) |  |
| Obesity | 1.019 (-2.666 to 4.705) |  |
| Comorbidity |  | 0.906 |
| None | Ref |  |
| One or more | 0.378 (-4.014 to 4.769) |  |
| Initial SARS-CoV-2 severity |  | 0.005 |
| Severe/critical | Ref |  |
| Mild/Moderate | 10.508 (3.217 to 17.8) |  |
| Symptoms | -1.705 (-2.595 to -0.816) | <0.001 |
| Days^c^ | -0.013 (-0.031 to 0.005) | 0.326 |
| CI: Confidence Interval. QoL was measured using the SF-36 questionnaire and Quality of life (QoL): was measured using SF-36 scores ranging from 0 (worst) to 100 (best). Clinical severity groups are defined as follows: Mild/moderate SARS-CoV-2 infection was defined as participants who reported not having had hospital admissions and oxygen therapy due to COVID-19 as well as home-based care and oxygen therapy and severe otherwise. Severe/critical SARS-CoV-2 infection was defined as participants reported having had hospital admission and/or needed oxygen therapy due to COVID-19.  The median time between a positive PCR test and enrolment into the study was 99 days (IQR= 71-169) and 38.8% (n=113) out of 291 participants had less than 90 days between a positive PCR test and first visit. | | |

**Identification**

**Screening**

**Eligibility**

**Included**

2145 patients screened for inclusion

1835 patients tested for COVID-19 at KUTRRH

300 patients tested for COVID-19 at KEMRI

10 walk-in patients

Patients with SARS-CoV-2 PCR test from December 2020 to July 2022 were selected

1395 patients excluded

- PCR test negative
- Clinically diagnosed
- Live outside Nairobi Metropolitan area

750 patients were invited to participate

459 patients refused to participate

291 patients included in the study

**Supplementary Figure S1:** Flow chart of participant enrolment into the cohort study.

Percentage of participants with symptom (%)

|  | Mild/Moderate | Severe/ Critical | Overall |
| --- | --- | --- | --- |
| Sore eyes | 11.7 | 11.9 | 11.7 |
| Upset Stomach | 11.7 | 16.7 | 12.4 |
| Weight loss | 11.2 | 31 | 14.1 |
| Wheeziness | 12.9 | 26.2 | 14.8 |
| Nausea | 14.1 | 31 | 16.5 |
| Dizziness | 19.3 | 28.6 | 20.6 |
| Sleep difficulties | 18.5 | 38 | 21.3 |
| Breathlessness | 20.5 | 40.5 | 23.4 |
| Stiff joints | 19.7 | 47.6 | 23.7 |
| Loss of strength | 28.5 | 50 | 31.6 |
| Headaches | 35.7 | 40.5 | 36.4 |
| Sore throat | 37.4 | 33.3 | 36.8 |
| Pain | 37 | 35.7 | 36.8 |
| Fatigue | 37.4 | 57.1 | 40.2 |
| Overall symptoms | 61 | 83.3 | 64.3 |

**Supplementary Figure S2:** Prevalence of COVID-19-related symptoms at the first study visit (x-axis). The median time between a positive PCR test and enrolment into the study was 99 days (IQR= 71-169) and 38.8% (n=113) out of 291 participants had less than 90 days between a positive PCR test and first visit.

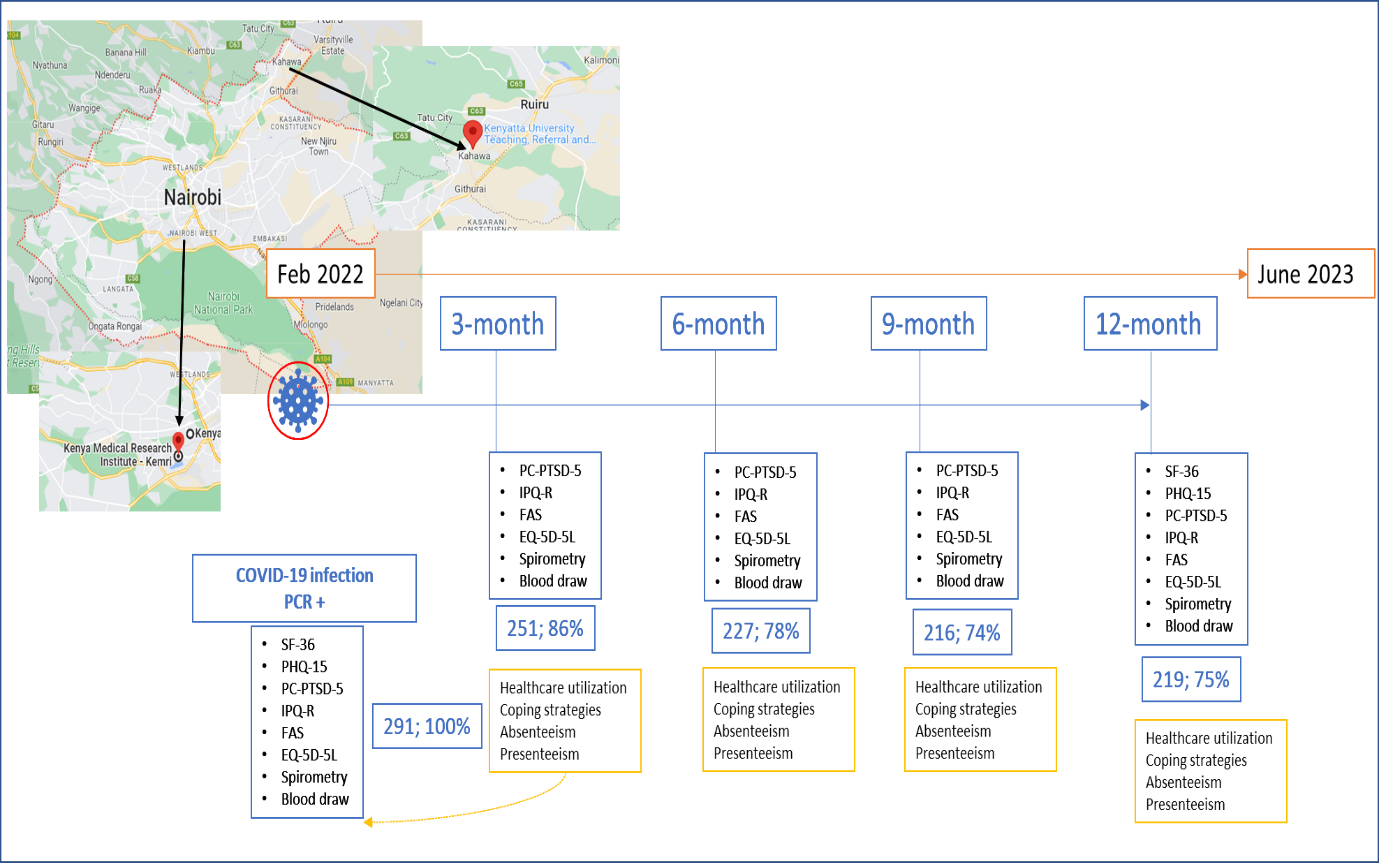

**Supplementary Figure S3: Overview of data collection in the Long COVID-19 study, Nairobi Kenya**

**Supplementary Figure S4. Transition between levels of self-reported severity of Long COVID over time among prospectively included participants, clinical severity group**

The vertical bars represent the number of participants for each severity level of the COVID-19-related symptom. Initial COVID-19 clinical severity groups are defined as follows: Mild/moderate SARS-CoV-2 infection was defined as participants who reported not having had hospital admissions and oxygen therapy due to COVID-19 as well as home-based care and oxygen therapy and severe otherwise. Severe/critical SARS-CoV-2 infection was defined as participants reported having had hospital admission and/or needed oxygen therapy due to COVID-19. The size of the streams indicates the number of study participants who moved from one severity level to another; transitioning from any severity level to “No” represents recovery from that symptom.

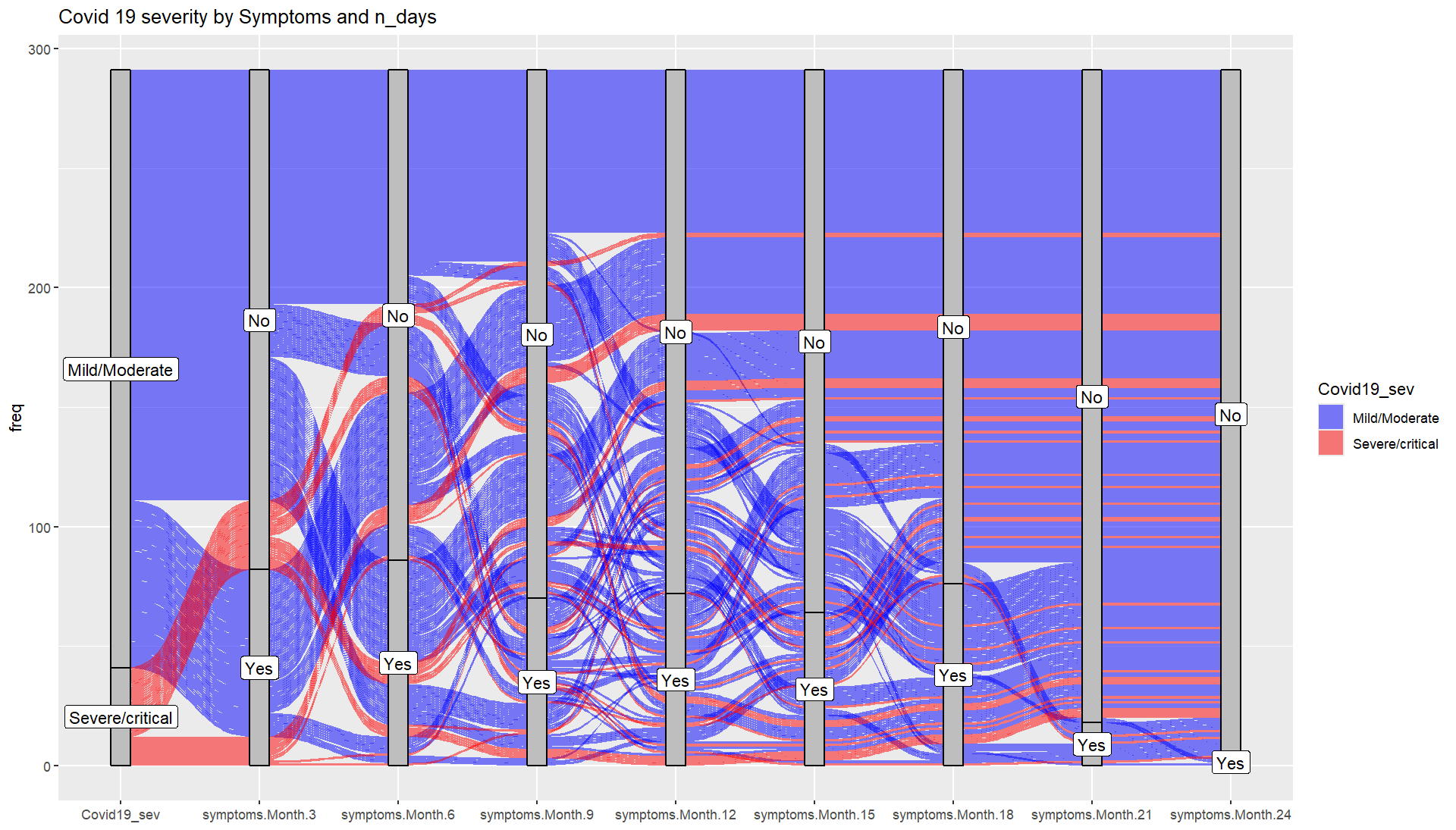

**Supplementary methods**

**Supplementary Methods 1: Illness Perception Questionnaire (IPQ-R)**

Your views about your illness

Listed below are a number of symptoms that you may or may not have experienced since your lung disease. Please indicate by circling Yes or No, whether you have experienced any of these symptoms since your COVID-19 disease, and whether you believe that these symptoms are related to your COVID-19 disease.

**This symptom is *related to my COVID-19 disease***

**I have experienced this symptom *since my COVID-19 disease***

Pain	Yes	No ________________ Yes	No

Sore Throat	Yes	No ________________ Yes	No

Nausea	Yes	No ________________ Yes	No

Breathlessness	Yes	No ________________ Yes	No

Weight Loss	Yes	No ________________ Yes	No

Weight Gain	Yes	No ________________ Yes	No

Fatigue	Yes	No ________________ Yes	No

Stiff Joints	Yes	No ________________ Yes	No

Sore Eyes	Yes	No ________________ Yes	No

Wheeziness	Yes	No ________________ Yes	No

Headaches	Yes	No ________________ Yes	No

Upset Stomach	Yes	No ________________ Yes	No

Sleep Difficulties	Yes	No ________________ Yes	No

Dizziness	Yes	No ________________ Yes	No

Loss of Strength	Yes	No ________________ Yes	No

**Supplementary methods 2: Test of the proportional-hazards assumptions after stcox using stcoxkm**

To verify the proportional hazards assumption in our Cox proportional hazards model, we used the stcoxkm command. This method compares the observed survival curves with the predicted survival curves under proportional hazards. Our analysis indicates that the proportional hazards assumption for COVID-19-related severity has not been violated where the observed and predicted values are close together. This suggests that the relationship between the covariates and the hazard of COVID-19-related severity remains consistent over time, validating the use of the Cox model in the study.

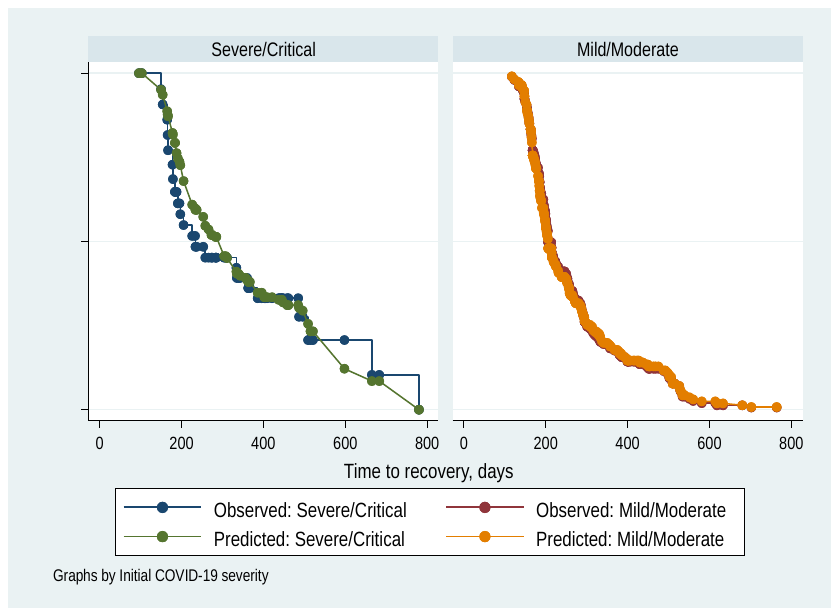
